# Absolute Benefit of Androgen Deprivation Therapy With Radiotherapy for Localized Prostate Cancer

**DOI:** 10.64898/2026.09.10.26362669

**Authors:** Jessica Aldous, William C. Jackson, Elizabeth Chase, Elise Covert, Joseph Tang, Udit Singhal, Todd M. Morgan, Daniel E. Spratt, Robert T. Dess, Matthew J. Schipper

## Abstract

**Objectives:** To estimate the absolute benefit of adding and prolonging androgen deprivation therapy (ADT) to radiation therapy (RT) for men with localized prostate cancer by integrating cancer-specific risk, other-cause mortality (OCM) risk, and relative treatment efficacy.

**Subjects and Methods:** Individualized risks were estimated by integrating cause-specific hazard estimates from a validated staging system (STAR-CAP) with those from a validated non-cancer mortality model (OCCAM). ADT treatment efficacy was then incorporated using published hazard ratio estimates from the MARCAP meta-analysis. Model utility was illustrated using the Prostate, Lung, Colon, and Ovarian (PLCO) cancer screening trial cohort (n=5468 prostate cancer patients with complete covariates). The primary outcome was the estimated 10-year absolute risk reduction (ARR) in distant metastasis (DM) from adding short-term ADT (STADT) to RT or extending STADT to long term ADT (LTADT).

**Results:** Within PLCO, model estimated risk of DM at 10 years ranged from <1% to 56% under guideline concordant care and OCM risk ranged from 3% to 80% independent of treatment. For patients with NCCN unfavorable intermediate-risk (n=1814), adding STADT to RT reduced estimated 10 year DM risk by a median of 4%, but the ARR estimates for individuals ranged from <1% to almost 15%. Similar variation was seen within NCCN high-risk patients (n=1289) with a median ARR of 9% [Range: 0.4%-17.2%] when prolonging ADT treatment. Moreover, 25% of NCCN unfavorable intermediate-risk patients have less than a 2.4% ARR while 25% of NCCN high-risk patients experience less than a 5% ARR.

**Conclusion:** An integrated model accounting for both prostate cancer aggressiveness and comorbidities demonstrates substantial heterogeneity in the estimated absolute benefit of ADT within conventional risk groups. This approach provides individualized estimates to support treatment discussions; external validation is needed before clinical implementation.

## Introduction

For patients with localized prostate cancer treated with definitive radiation therapy (RT), the addition and prolongation of androgen deprivation therapy (ADT) reduces metastasis and improves disease-specific survival (1). However, localized prostate cancer is a heterogeneous disease with broad prognostic variability. This variability is highlighted by clinically significant differences in estimated 10 year risk of prostate cancer-specific mortality (PCSM) within the international staging collaboration for cancer of the prostate (STAR-CAP) clinical prognostic risk groupings (2), which range from 1% in Stage IA to greater than 20% in Stage III.

Despite this prognostic range, NCCN guidelines utilize a broad five-tiered disease risk categorization (low, favorable intermediate, unfavorable intermediate, high, and very high). Consequently, most eligible patients with high-risk disease are recommended long term ADT (LTADT), and those with unfavorable intermediate-risk disease are recommended short-term ADT (STADT), regardless of the absolute magnitude of the treatment benefit they are likely to receive. Furthermore, while guidelines suggest incorporating life expectancy into decision-making, validated, easy-to-use tools to estimate other-cause mortality (OCM) have been lacking (3,4).

Accurate OCM estimation is critical given that more than 50% of men treated with curative RT are over the age of 65, a demographic enriched for high competing risks of mortality (5,6). To address this, we previously developed and validated the Other-Cause Comorbidity-Adjusted Mortality (OCCAM) model which includes eight predictors: age; education; marital status; diabetes; hypertension; stroke; body mass index; and smoking status (3).

Given the significant heterogeneity in both individual patient risk of PCSM and OCM, we propose a model that integrates the STAR-CAP model with our OCCAM model and ADT benefit hazard ratios (HR) from a MARCAP meta-analysis (1). This integrated approach estimates absolute risk reduction (ARR) in distant metastasis (DM) and PCSM for individual patients with differing comorbidities under three potential treatment strategies: RT alone (RT), RT with STADT, and RT with LTADT. We hypothesized that using this approach would reveal significant variability in ARR estimates both within and across standard NCCN risk groupings.

## Patients and Methods

### Patients

We used two datasets to construct and illustrate our model. The STAR-CAP dataset was used to construct the integrated model’s disease risk estimates (7). The second dataset, which illustrated the utility of the model, was a subgroup of 5,468 patients 1) treated with radical prostatectomy/radiation and 2) non-missing OCCAM and STAR-CAP variables from the cohort of 8,220 patients in the Prostate, Lung, Colon and Ovarian (PLCO) cancer screening trial diagnosed with non-metastatic prostate cancer (3). General characteristics of patients in STAR-CAP and PLCO are in Supplement Table S2. This study was reported in accordance with TRIPOD (7) and with consideration of the PATH consensus criteria (8) (See Supplement Table S4 and S5).

### Development of the Prognostic Model Integrating STAR-CAP and OCCAM

We used a standard method to integrate patient-specific models for two competing risk events (9). Specifically, we combined the cause-specific hazard of dying from prostate cancer (λ_1_) derived from STAR-CAP with the cause-specific hazard of dying from other causes (λ_2_) derived from OCCAM to calculate the absolute risk of DM or PCSM conditional on the combined set of predictors. We estimated λ_1_ in the STAR-CAP data set from a Cox model with stage as a nine-level categorical covariate and λ_2_using the previously published OCCAM. Detailed statistical methodology is provided in the Supplement.

### Incorporating Treatment Effect of ADT Use and Duration

We incorporated treatment effects to provide PCSM estimates individualized by treatment, together with OCM estimates, for three strategies: RT alone, RT plus STADT, and RT plus LTADT. We used hazard ratios from relevant randomized trials from the MARCAP meta-analysis in which the HRs for RT + STADT vs. RT alone on DM and PCSM are 0.64 and 0.55, respectively, while the HRs for RT + LTADT vs. RT + STADT on DM and PCSM are 0.65 and 0.59, respectively (1).

To estimate the cause-specific hazard of PCSM as a function of STAR-CAP and treatment, we treated observed stage-specific survival as a weighted average of the treatment and stage-specific survival curves. The weights were calculated as the proportion of patients in the STAR-CAP dataset that received each treatment. We constrained these estimates so that the resulting ADT treatment effects equal the published HRs stated previously (details in Supplement). Solving this system of equations allows us to estimate (a) DM/PCSM hazard under each of the three treatments (b) at specific time points (6-month intervals over 20 years following treatment), and (c) for each STAR-CAP stage. These treatment specific hazards are used as λ_1_ in our integrated model, allowing us to calculate the PCSM risk accounting for OCM risk (Supplement). We used the same approach to estimate cumulative incidence of DM and summed the DM and OCM hazards to derive metastasis-free survival (MFS) probability. Critically, we assume there is no treatment effect on OCM, though our model could easily be modified to allow for differential OCM risk by treatment as for example shown recently for LTADT (5). The resulting integrated model is available as an online clinical decision aid at micapmodels.org.

### Applying the Integrated Model to PLCO

To demonstrate the integrated model’s utility, we used the PLCO screening trial prostate cohort, which includes the demographic and comorbidity information needed to generate integrated predictions. We limited the PLCO cohort to men diagnosed with prostate cancer and who underwent radical prostatectomy or radiation therapy as primary treatment. Because PLCO is an observational study with treatment confounding and missing variables (percent positive cores, Gleason 3+4 vs Gleason 4+3), we used a simulation approach.

We created standardized patient profiles by mapping each PLCO patient’s observed OCM profile to various STAR-CAP stages. This allowed us to use the integrated model to calculate the ARR of DM, PCSM, and OCM for an observed distribution of OCM risk in a localized prostate cancer population and across different cancer stages. We then calculated treatment benefit for individual patients as DM or PCSM ARR at 10 years, comparing RT+STADT versus RT (for Stages IC-IIIB) and RT+LTADT versus RT+STADT (for Stages IIA-IIIC).

## Results

### Risk of Prostate Cancer Outcomes Depends on Other Cause Mortality Risk

The estimated risk of non-cancer mortality within 10 years of treatment, across PLCO patients, ranged from 3% to 80%. To illustrate the impact of the competing risk of OCM on prostate cancer outcomes, we modeled three representative patients from PLCO with OCM risk that is ‘low’ (5th percentile), ‘average’ (median) and ‘high’ (95th percentile). The average OCM risk patient is 74 years old, married, and has completed some college. He is a former smoker, overweight (BMI 27.1), and has a history of hypertension, but no diabetes nor previous stroke (see Supplement Table S3 for characteristics of the low and high-risk patients). Figures 1A and S1A display the cumulative incidence curves for DM and PCSM over 15 years for the three representative patients all with STAR-CAP Stage IIC treated with RT+STADT. Although these men have identical prostate cancer clinical characteristics and treatment, their predicted outcome trajectories differ substantially. As OCM increases, the absolute risk of DM and PCSM will necessarily decrease due to increasing competing risk. At 10 years, the low OCM risk patient has absolute risk estimates of 16% and 8% for DM and PCSM, respectively. In contrast, the high OCM risk patient has absolute risk estimates of 12% and 5% for DM and PCSM, respectively. By way of context, this 3% absolute difference in PCSM risk is greater than the increase in risk when moving from STAR-CAP Stage IIA to IIIA.

**Figure 1:**
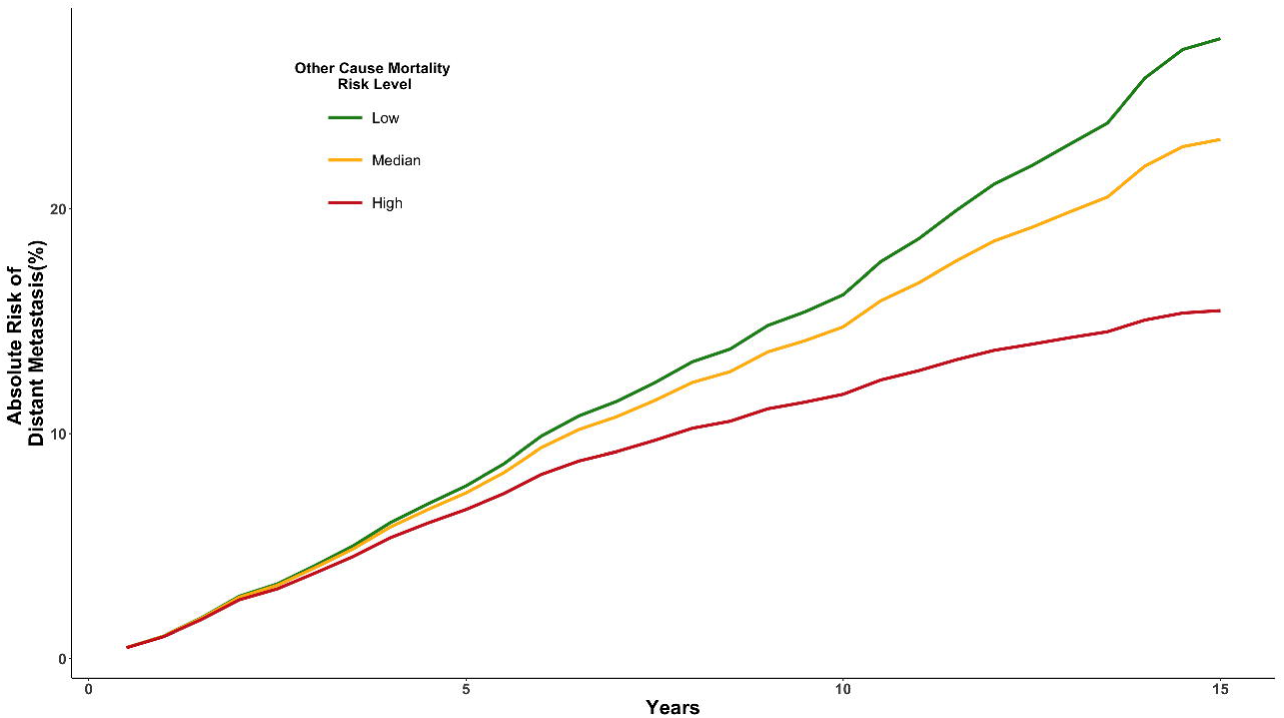
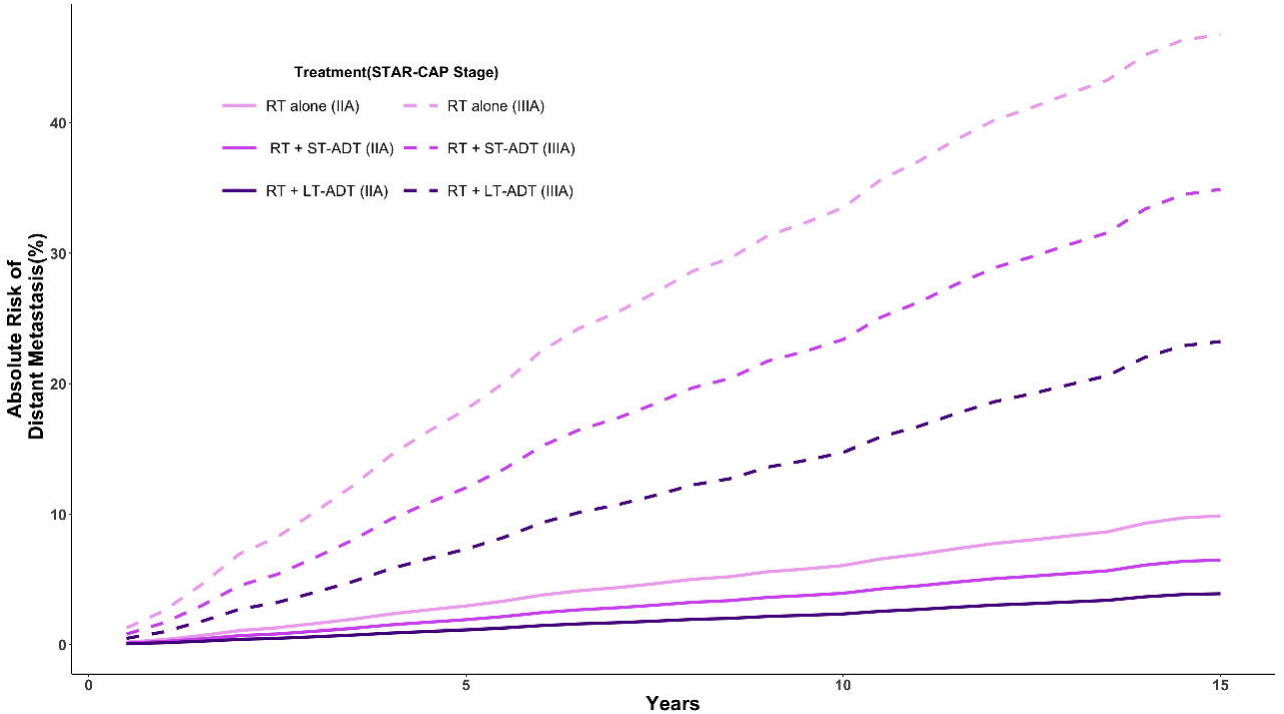
Impact of life expectancy (A) and treatment (B) on distant metastasis. **(A)** Each curve represents the absolute risk of experiencing distant metastasis **for a STAR-CAP IIC patient** across time for a given other cause mortality risk group. Life expectancy is represented by variable other cause mortality risk (OCM) within the PLCO cohort. Low risk is defined as the 5th percentile of OCM risk in PLCO patients (green). Median risk is the 50th percentile of OCM risk in PLCO patients (yellow). High-risk is the 95th percentile of OCM risk in PLCO patients (red). (B) Each curve represents the absolute risk of experiencing distant metastasis across time **for STAR-CAP IIA and IIIA patients** and treatment assignment. STAR-CAP stage is indicated by line type (IIA: solid; IIIA: dashed). Color distinguishes treatment with light purple for radiation alone, a medium shade for radiation + short term ADT, and the darkest shade for radiation + long term ADT.

### Individualized Predictions of DM and PCSM Depend on Treatment

Within PLCO, model estimated risk of DM at 10 years ranged from <1% to 56% under guideline-concordant care. Figure 1B compares the absolute risk of DM for an average OCM risk patient between STAR-CAP stage IIA and IIIA patients over time. The absolute risk of DM over 10 years under RT alone, RT+STADT or RT+LTADT are 6%, 4% and 2% for the IIA patient and 34%, 23%, and 15% for the IIIA patient (PCSM differences are in Figure S1B). Consequently, the treatment benefit of adding ADT to the IIA patient is only around a 2% reduction in 10 year absolute risk of DM. By contrast, the benefit of extending ADT to the IIIA patient is greater; with approximately 8% reduction in absolute risk of DM at 10 years.

### Heterogeneity of Treatment Effect Within and Between NCCN Risk Groups

The distributions of patient-level estimated benefit of RT+STADT vs RT and RT+LTADT vs RT+STADT are shown in Figure 2 for unfavorable intermediate and high NCCN risk groups. Treatment benefit is quantified as the ARR of DM within 10 years. Notable variation is seen within each NCCN risk group. In patients with unfavorable intermediate-risk disease, the model estimates adding STADT to RT reduces the cumulative incidence of DM by approximately 4% for the average patient, but the benefit ranges from less than 1% to almost 15%; 25% of patients derive less than 2.4% benefit from adding ADT. Within high-risk patients, the median estimated benefit of LTADT over STADT is approximately 9% but ranges from <1% to 17%. Approximately 25% of patients obtain less than a 5% ARR in DM at 10 years (Figure 2). Figure S2 shows similar results for treatment effect on PCSM.

**Figure 2:**
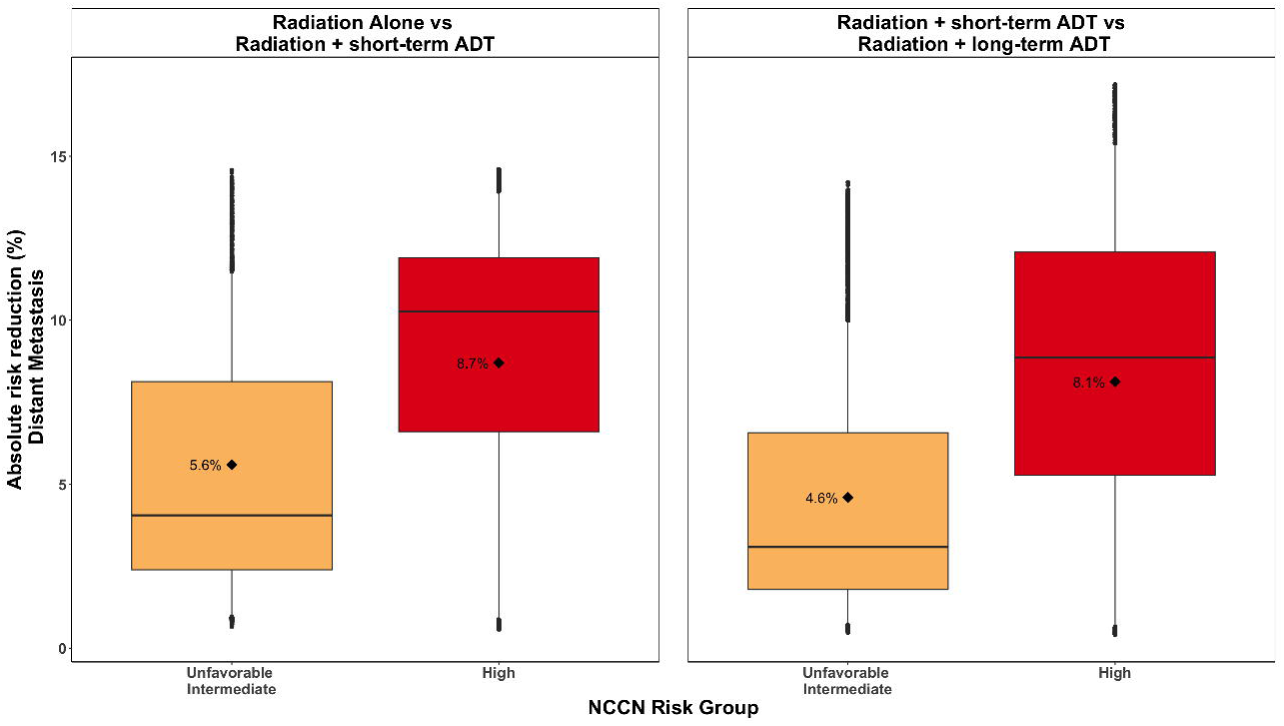
Variation in individual patient level estimates of treatment benefit in distant metastasis from addition of STADT to RT and LTADT to STADT across patients within the NCCN unfavorable intermediate and high-risk groups. Treatment benefit is defined as the absolute reduction in distant metastasis risk when adding ADT to radiation (left) or prolonging ADT treatment (right). The bottom and top edges of the box indicate the 25th and 75th percentiles of absolute risk reduction, respectively. The bold middle line identifies the 50th percentile and the whiskers extend to the 5th and 95th percentiles of absolute risk reduction. The mean value is labeled by a diamond and its numeric percentage. Variability in treatment benefit within- and between-NCCN unfavorable intermediate (orange) and high (red) risk groups is attributable to individual differences in STAR-CAP staging and other cause mortality risk.

In addition to the variation of benefit within NCCN risk group, there is substantial overlap in benefit between UIR and high-risk. For example, if LTADT were to be recommended for patients likely to derive an ARR benefit of at least 5%, then more than 25% of UIR patients would meet these criteria while more than 25% of high-risk patients would not meet it.

Figure 3 illustrates that these ADT treatment benefits vary with OCM risk, even within a STAR-CAP stage. At 10 years, STAR-CAP stage IIA patients with low OCM risk (green curves) experience an additional estimated ARR of 0.6% in DM risk compared to their high OCM risk counterparts (red curves) when adding STADT to RT (2.3% vs 1.7%). Similarly, STAR-CAP stage IIIA patients with low OCM risk have a 9% estimated ARR in DM at 10 years when extending ADT compared to only a 7% reduction for IIIA patients with high OCM risk profiles.

**Figure 3:**
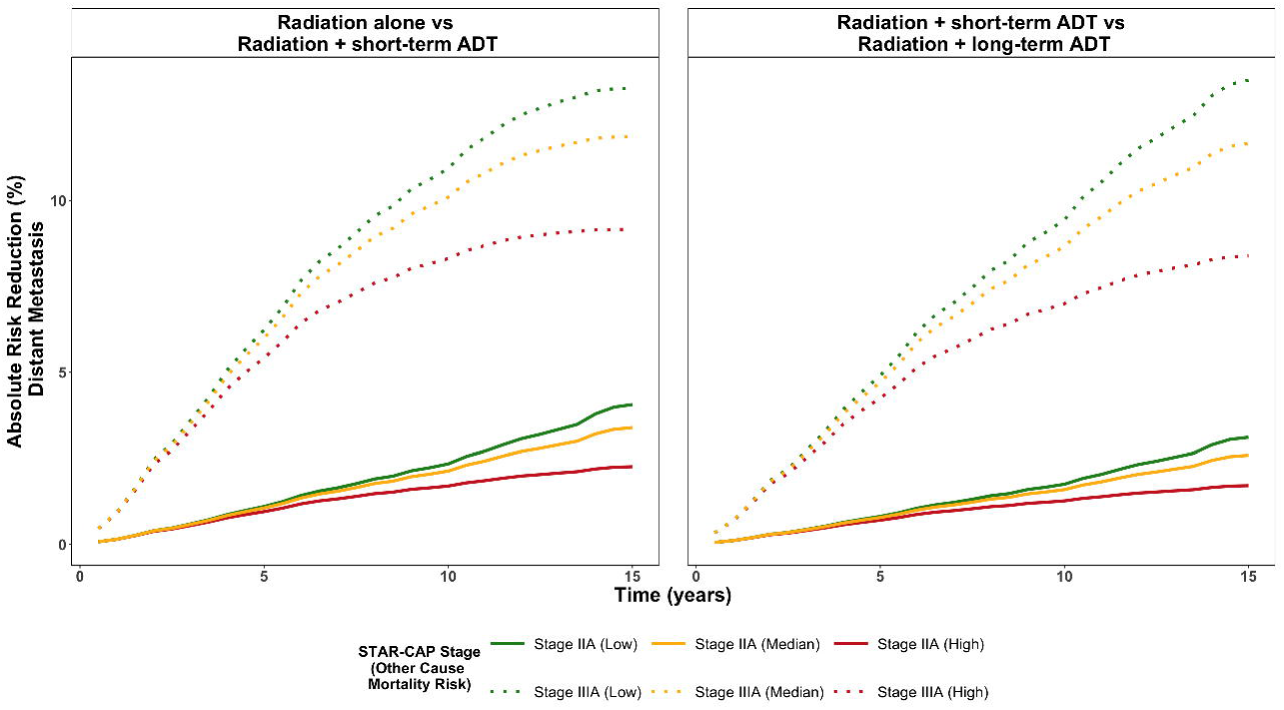
Treatment benefit in distant metastasis over time for STAR-CAP IIA and IIIA patients, colored by other cause mortality risk levels. Treatment benefit is defined as the absolute reduction in distant metastasis risk at a given time when adding ADT to radiation (left) or prolonging ADT treatment (right). Low risk is defined as the 5th percentile of other cause mortality risk (OCM) risk in PLCO patients (green). Median risk is the 50th percentile of OCM risk in PLCO patients (yellow). High-risk is the 95th percentile of OCM risk in PLCO patients (red). STAR-CAP stage is indicated by line type (IIA: solid; IIIA: dashed).

### Integrated Estimates

Table 1 shows the 10-year estimates presented on the integrated app for a patient with STAR-CAP stage IIC and median OCM risk. Figure S4 shows an app-derived pictogram visually representing the estimated DM (4A) and PCSM (4B) risk compared to OCM risk estimates for said patient to facilitate patient understanding(10,11).

## Discussion

While national guidelines recommend fixed durations of ADT with curative RT for each NCCN risk category, this one-size-fits-all approach does not account for the significant interplay between tumor aggressiveness and competing mortality risk. Our analysis demonstrates that guideline concordant care may result in overtreatment for a subset of men with high-risk disease who, due to competing OCM risk, derive small absolute benefits from the addition or prolongation of ADT. Conversely, specific patients with unfavorable intermediate-risk disease may experience greater benefit from treatment intensification than classical risk stratification may suggest. By integrating independently validated models for cancer-specific (STAR-CAP) and other-cause mortality (OCCAM), our freely available model provides quantitative estimates that may help shift clinical decision making from algorithmic assignment based on risk group to personalized recommendations based on expected treatment benefits.

Our findings are comparable to a secondary analysis of NRG/RTOG 9408 assessing whether stratifying patients by their risk of competing mortality using generalized competing event modeling could selectively identify patients that benefitted from the addition of STADT to RT (12). They fit a new model incorporating many of the same variables (e.g. Gleason, PSA, age, comorbidities). Utilizing different statistical methods, they also found that increasing Gleason grade, lower age and lack of any comorbidity were associated with higher ‘omega’ scores which were associated with greater ADT benefit. Notable differences between this analysis and ours include use of randomized data in the NRG/RTOG 9408 analysis. However, their model lacks external validation whereas we used two independently developed and externally validated models for both cancer outcomes (STAR-CAP) and competing risk (OCCAM). Additionally, we estimated treatment benefit as differences in the absolute risk of DM/PCSM at 10 and 15 years rather than as hazard ratios. Despite these differences, both analyses support the notion that the absolute benefit of the addition of ADT to RT is dependent on risk of both PCSM and OCM.

Limitations of our results include that our models do not currently incorporate clinically available prognostic/predictive biomarkers such as the 22-gene Decipher genomic classifier or the multimodal artificial intelligence test (ArteraAI) (13,14). However, the modular framework presented here allows for validated prognostic / predictive biomarkers to be incorporated within our integrated model in the future. Additionally, our analyses include data from patients treated dating back to the early 1990s. Since this time, there has been significant grade and stage migration, most notably with the 2005 (15) and 2014 (16) International Society of Urological Pathology changes to the Gleason scoring system, and more recently with the incorporation of molecular imaging (i.e. Prostate-Specific Membrane Antigen PET) for pre-treatment staging, and these changes may result in biased treatment benefit estimates in our analyses. Lastly, our statistical approach makes several assumptions, namely: the hazards of DM/PCSM and OCM are conditionally independent, ADT HRs do not vary with patient covariates, and ADT use/duration does not negatively impact OCM. This highlights the need for external validation in randomized data. Although the two component models have been externally validated, it will be important to separately validate the patient-level predicted treatment benefit from the integrated model. Validation in NRG/RTOG 9202 (17), 9408 (18), and 0815 (19) is ongoing and will be reported separately in the future. In addition to these validation efforts, future work will focus on updating models to include relevant predictive / prognostic biomarkers and identifying patients who may be under or over treated with ADT.

In conclusion, we provide an integrated model which incorporates an individual patient’s risk of PCSM and OCM to provide personalized absolute benefits of ADT (short and long term) in addition to RT in patients with localized prostate cancer. This model and the accompanying software fill a current gap in individualized care for patients with localized prostate cancer, providing quantitative risk estimates to facilitate nuanced treatment discussions between clinicians and their patients. External validation of patient-level benefit is needed to broaden clinical implementation.

## Data Availability

PLCO data is available upon request at https://cdas.cancer.gov/learn/plco/instructions/.
The cause-specific hazards used and the code to generate these estimates are available upon reasonable request to the authors.

## Acknowledgements

The work of J.A. and E.C. was partially supported by National Institutes of Health grant CA83654.

---

**Table 1: Example risk at 10 years for a STAR-CAP IIC patient with median OCM risk available through the app.** The table presents the 10 year absolute risk estimates of four outcomes of interest across treatments.

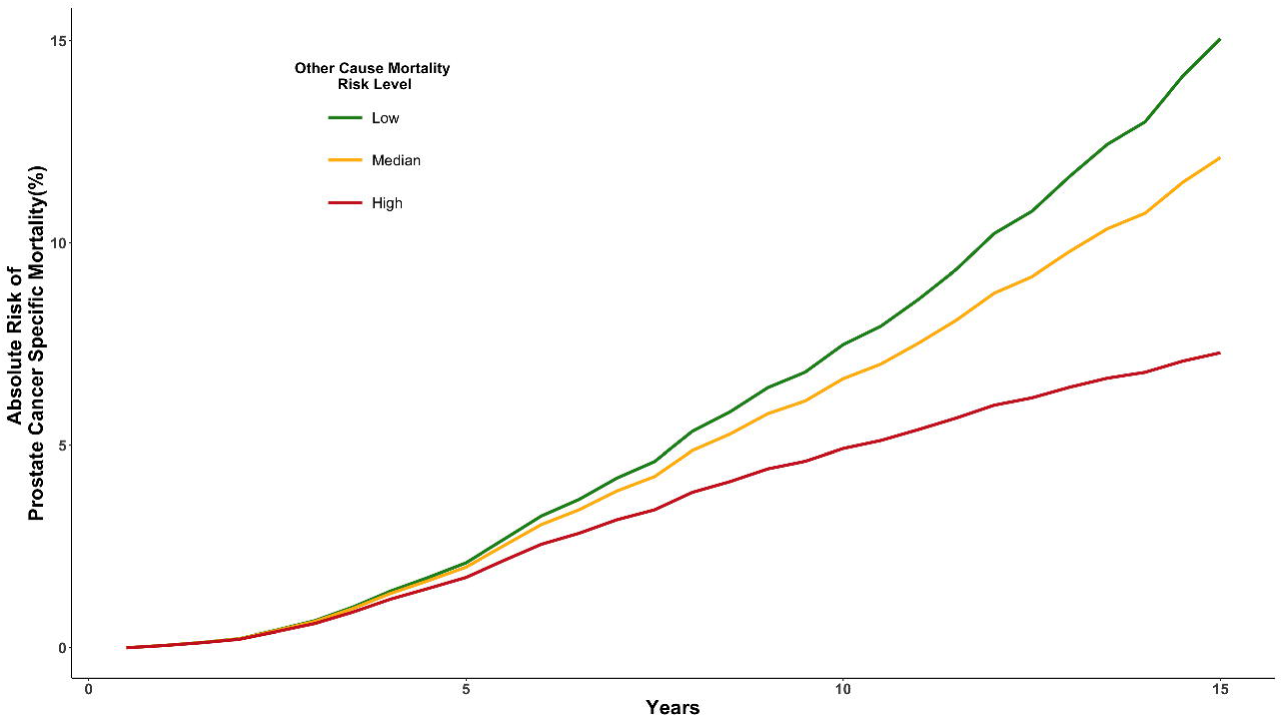

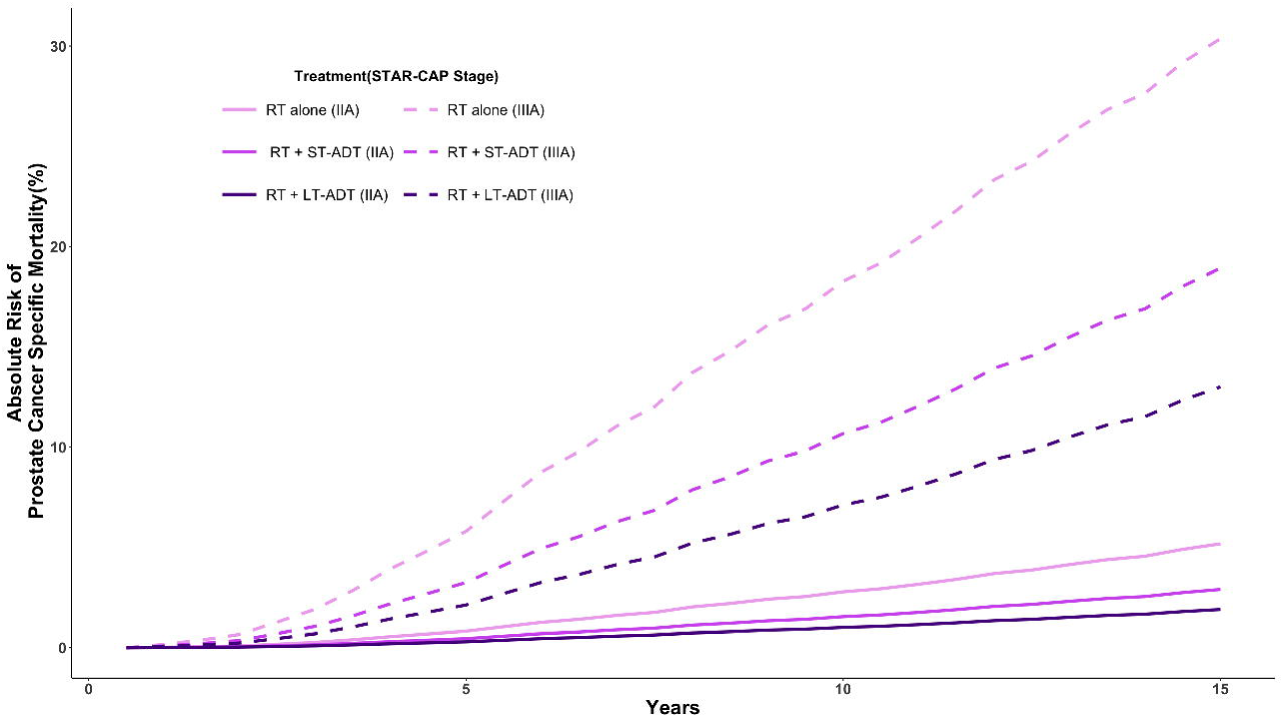

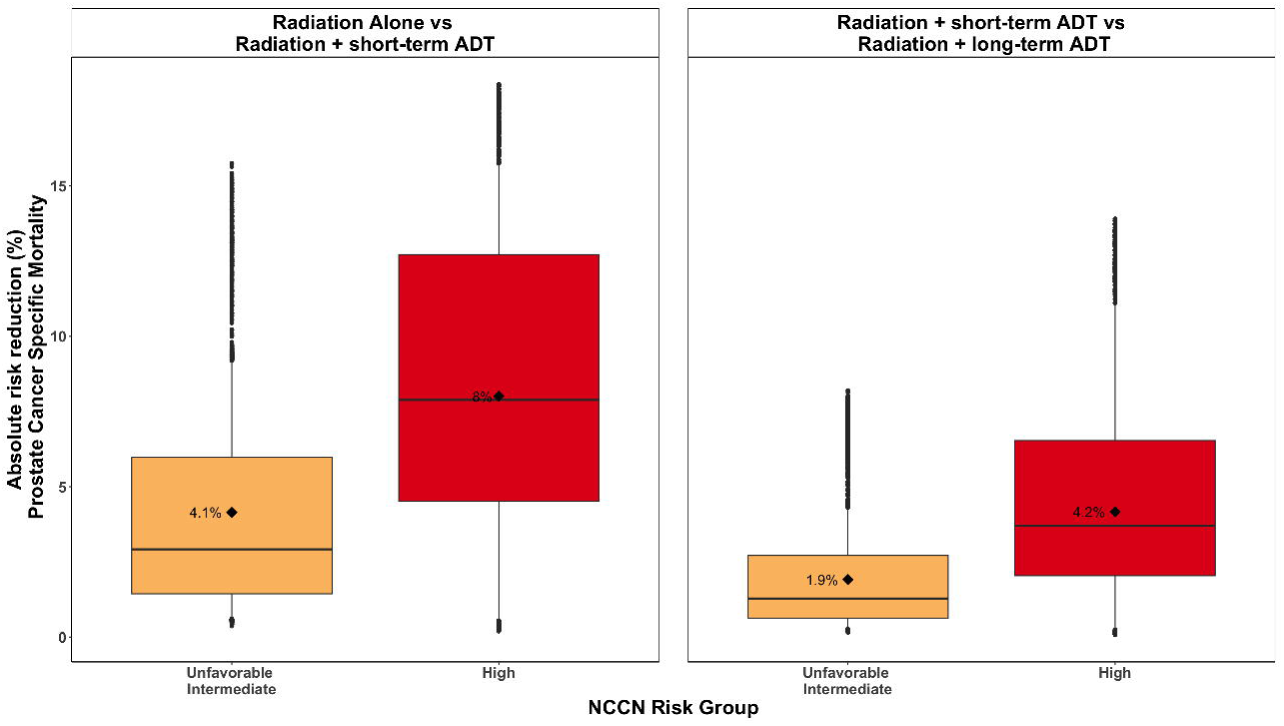

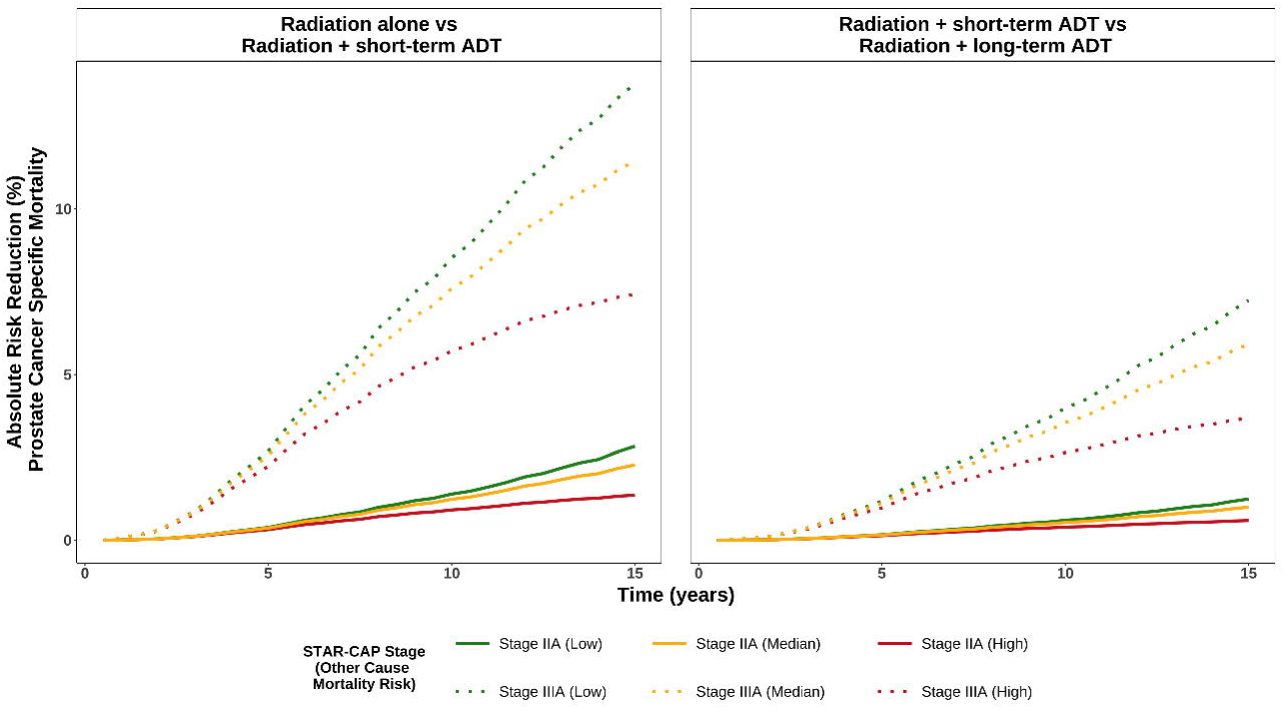

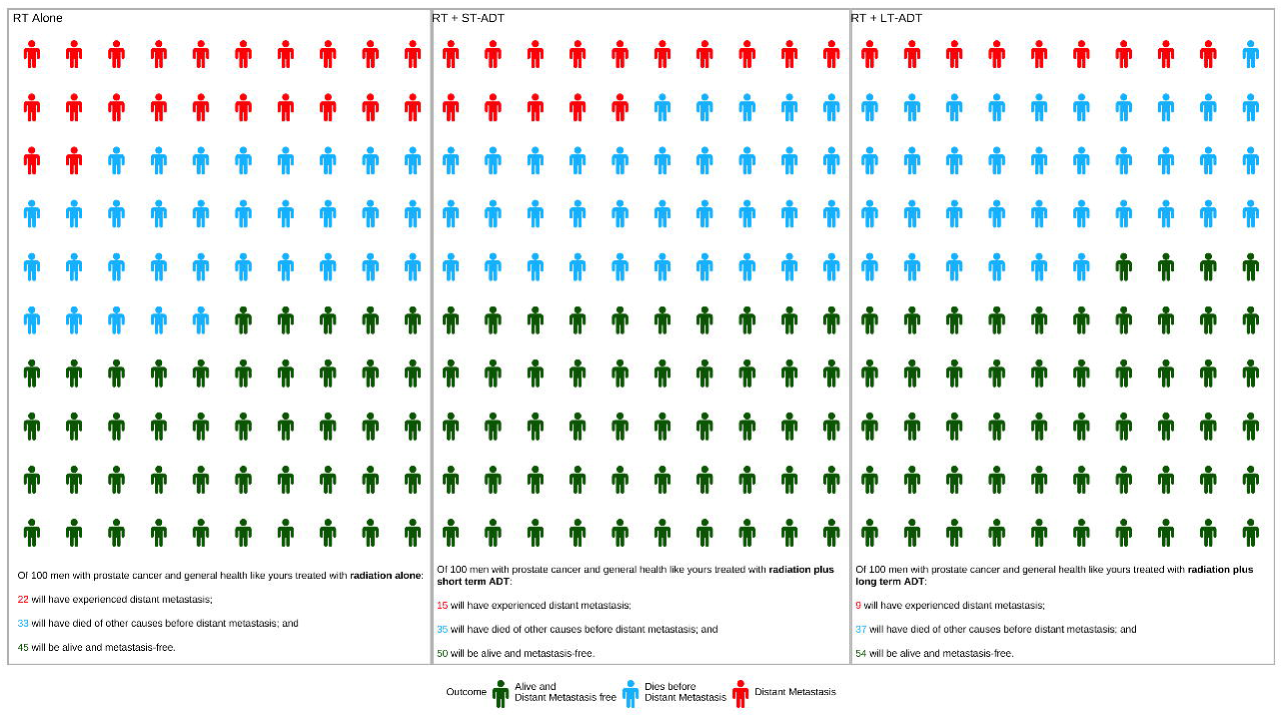

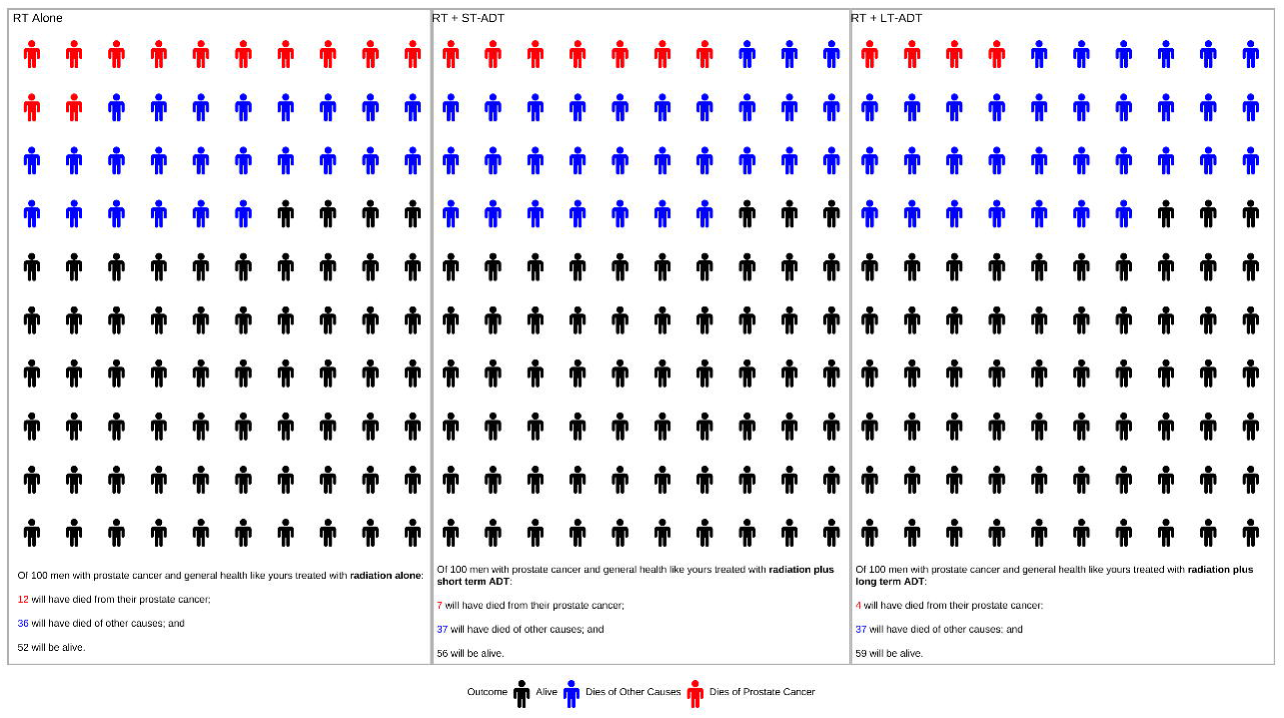

