## Supplement Material for "Absolute Benefit of Androgen Deprivation Therapy With Radiotherapy for Localized Prostate Cancer"

### Data Sources Demographics and Limitations

PLCO is the oldest of the three cohorts with median age 69 (IQR: 65, 72), followed by STAR-CAP (64; IQR: [59, 70]) and then NHANES (60; IQR: [50, 70]). PLCO is also the least diverse of the three cohorts, with 86.4% of patients identifying as White, compared to 62.7% and 51.6% of STAR-CAP and NHANES patients, respectively. Overall, PLCO patients are healthier than NHANES patients, despite being older, and have characteristics of more affluent patients. We also note that NHANES is not a prostate cancer patient population. Only 5.1% of NHANES patients had been diagnosed with prostate cancer at the time of data collection. In terms of cancer characteristics, PLCO patients generally had more advanced cancer than STAR-CAP patients. They were less likely to receive radical prostatectomy as primary treatment, instead receiving RT at higher rates. Rates of receiving adjuvant hormonal therapy were similar among radiation patients in both cohorts, at about 50%. PLCO does not provide information on the timing of adjuvant hormonal therapy, so we are unable to compare the distribution of timing between the two cohorts. See Table S1 and S2 for additional details.

### Methods

The integrated model uses the predictors of both STAR-CAP and OCCAM. Using a points-based system, STAR-CAP assigns patients to one of nine stages (IA-IIIC), created by adding up point contributions from age, clinical T stage, clinical N stage, Gleason grade, percent positive cores, and prostate specific antigen (PSA) level. These nine stages are used as a categorical predictor in the STAR-CAP model; each stage has a corresponding risk trajectory. OCCAM relies on eight predictors: age, body mass index (BMI), educational attainment, marital status, smoking status, and previous diagnosis of diabetes, hypertension, or stroke.

The integrated model considers outcomes from three first-line definitive radiation-based courses of treatment: radiation therapy (RT) alone, RT plus short-term androgen deprivation therapy (STADT), and RT plus long term androgen therapy (LTADT). In the STAR-CAP cohort, short-term ADT is defined as a duration less than 12 months. Long term ADT has a duration of 12 months or greater. Radiation therapy includes both external beam radiation therapy and brachytherapy. While the STAR-CAP cohort does include men who underwent radical prostatectomy, the comparison of surgery versus radiation-based treatment outcomes is left for future research. The handling of surgery patients in computation is described in further detail below.

The primary endpoints were time to distant metastasis (DM), time to death of other causes (OCM), and time to death of prostate cancer (PCSM). Time to DM was defined as the time from treatment initiation to detection of metastatic disease via imaging. Time to OCM was defined as the time from treatment initiation to death due to causes excluding prostate cancer. Time to PCSM was defined as the time from treatment initiation to death due to prostate cancer censored at last follow-up. In STAR-CAP, death due to prostate cancer was defined per individual institution, but most often required metastatic and/or castration-resistant cancer present at the time of death without alternative cause. In STAR-CAP, imaging for metastasis was per institution, usually motivated by symptoms or continued increase in PSA levels.

Hazard estimates in the original STAR-CAP paper^2^ were estimated using Fine-Gray modeling. To enable model integration for the purposes of this project, we re-fit a Cox proportional hazards model to estimate the cause-specific hazard of PCSM over time by categorical STAR-CAP stage. The same was done for the outcome of DM. We fit the cause-specific models using the full STAR-CAP cohort, where each patient was assigned a stage (IA-IIIC) based on the points-based system described by Dess et al. (2020). OCCAM provides cause-specific hazard estimates; these were outputted directly from the OCCAM model (3).

Let $\lambda_{1}(t|z_{1})$denote the cause-specific hazard of dying from prostate cancer at time t conditional on predictors $z_{1}$, and $\lambda_{2}(t|z_{2})$denote the cause-specific hazard of dying of other causes at time t conditional on predictors $z_{2}$. Define $r_{1}(t|Z)$as the absolute risk of dying of prostate cancer at time t, conditional on predictors Z, which is the combined set of all predictors across all causes. Estimates of absolute risk of PCSM can be obtained according to the following formula (9):

$$r_{1}\left( t | Z \right)= \int_{0}^{t} \lambda_{1}(u|z_{1})exp(-\int_{0}^{u} \lambda_{1}\left( s | z_{1} \right)+\lambda_{2}(s|z_{2})ds)du$$

We estimated $\lambda_{1}$in the STAR-CAP dataset from a Cox model with stage as a 9-level categorical covariate, using the Nelson-Aalen estimate for the baseline cumulative hazard. We estimated $\lambda_{2}$using the previously published OCCAM model.

A key assumption we make here is that the HRs for the effect of treatment on PCSM and DM are constant and do not vary across patient covariates (20). Additionally, we set the HR for the effect of treatment on OCM to be 1.0. Surgery patients were handled in a stage-specific manner, with the proportion of patients in a given stage who were treated with radical prostatectomy grouped into the weight for the radiation-based treatment considered optimal for that stage. The optimal radiation-based regimen per stage was decided based on clinician input. Surgery was grouped with RT alone for stages IA-IC, with RT plus STADT for stages IIA-IIB, and with RT plus LTADT for stages IIC-IIIC. To solve for the unknown treatment-specific survival values we begin with three equations for each stage and at each time across a grid of timepoints and separately for PCSM and DM. In the first equation, the average (across treatments) survival is written as a weighted average of the treatment specific survivals $S_{TREATMENT}\left( t | STARCAP \right)$. The weights are calculated as the proportion of patients in that STAR-CAP stage who received that treatment. We relate the two ADT treatment specific survivals using published HRs from a meta-analysis of 10 phase 3 clinical trials. With this approach we have three equations to solve for the three unknowns (i.e. the survival for each of the 3 treatments).

$$S\left( t | STARCAP \right)= w_{RT}S_{RT}\left( t | STARCAP \right)+ w_{STADT}S_{STADT}\left( t | STARCAP \right)+w_{LTADT}S_{LTADT}\left( t | STARCAP \right)$$

$${S_{STADT}\left( t | STARCAP \right)= S}_{RT}\left( t | STARCAP \right)^{HR_{RTvsSTADT}}$$

$${S_{LTADT}\left( t | STARCAP \right)= S}_{STADT}\left( t | STARCAP \right)^{HR_{STADTvsLTADT}}$$

An identical process was repeated for the outcome of DM.

**Supplemental Tables and Figures**

Table S1: Data sources used in integrated model and treatment benefit prediction.

Table S2: Characteristics of the STAR-CAP and PLCO cohorts, which were used to build and illustrate the integrated model.

Table S3: Comorbidity profiles for low, median, and high OCM risk patients from the PLCO prostate cancer cohort.

Table S4: TRIPOD Guidelines Checklist.

Table S5: The Predictive Approaches to Treatment effect Heterogeneity (PATH) Consensus Criteria.

Figure S1: Impact of life expectancy(A) and treatment(B) on prostate cancer specific mortality. (A) compares mortality within for a STAR-CAP IIC patient across three other cause mortality risk groups while (B) compares absolute risk of mortality over time for STAR-CAP IIA and IIIA patients, under three potential treatments.

Figure S2: Variation in individual patient level estimates of treatment benefit in prostate cancer specific mortality from addition of STADT to RT and LTADT to STADT across patients within the NCCN unfavorable intermediate and high-risk groups.

Figure S3: Treatment benefit in prostate cancer specific mortality over time for STAR-CAP IIA and IIIA patients, colored by other cause mortality risk levels.

Figure S4: Example of distant metastasis (A) and prostate cancer specific mortality (B) pictograms at 10 years for a STAR-CAP IIC patient with median OCM risk available through the app

### Table S1: Data sources used in integrated model and treatment benefit prediction.

| **Component** | **Description** | **Training Data** | **Validation Data** |
| --- | --- | --- | --- |
| Prostate cancer staging model | STAR-CAP is a pretreatment clinical prognostic stage group system for nonmetastatic prostate cancer developed using a large, diverse international cohort treated with standard curative treatment options. The proposed AJCC-compliant clinical prognostic stage group systems for prostate cancer outperformed the existing AJCC system and commonly used risk-stratification systems (2). | STAR-CAP training data | - STAR-CAP validation data - SEER - Retrospective data from RP-treated prostate cancer patients in the single-institution Martini-Klinik database (21) |
| Cause-specific OCM model | Using two national cohorts, Chase et al. ([2022](https://bjui-journals.onlinelibrary.wiley.com/doi/abs/10.1111/bju.15740)) developed and validated a prediction model for other-cause mortality for patients with prostate cancer treated in the United States. The final model included 8 predictors: age, education level, marital status, diabetes, hypertension, stroke, BMI, and smoking status. | NHANES, restricted to patients:   - Male - Over age 40 - Free of non-prostate malignancy - With complete data for OCCAM predictors | PLCO, restricted to patients:   - Diagnosed with prostate cancer - With complete data for OCCAM predictors   We further restrict to patients who underwent RP or RT for integrated model illustration. |
| Treatment HRs | Individual patient data was obtained from 10 radiotherapy-related Phase III RCTs and used in a meta-analysis to evaluate prognostic and predictive performance of standard clinicopathologic variables. Endpoints included distant metastasis (DM), metastasis-free survival (MFS), overall survival (OS), and prostate cancer specific mortality (PCSM). Analysis was conducted separately for RT alone versus RT + STADT and RT + STADT versus RT + LTADT. (1,22) | - NRG/RTOG 9202, 9408, 9413, 9910, 0126 - EORTC 22863, 22961, 22991 - DART 01/05 GICOR - Ottawa 0101 |  |

**Table S2: Characteristics of the STAR-CAP and PLCO prostate cohort who received radiation or radical prostatectomy, which were used to build and illustrate the integrated model.**

| **Characteristic** | **STAR-CAP** | **PLCO** |
| --- | --- | --- |
| **N** | 19684 | 5468 |
| **Age at diagnosis** |  |  |
| Median (IQR) | 64.0 (59.0, 70.0) | 68.0 (65.0, 72.0) |
| **Race (%)** |  |  |
| White | 12333 (62.7) | 4874 (89.1) |
| Black | 2138 (10.9) | 296 (5.4) |
| Other | 664 (3.4) | 296 (5.2) |
| Unknown | 4549 (23.1) | 2 (0) |
| **Prostate cancer (%)** | 19684 (100.0) | 5468 (100.0) |
| **Clinical tumor category (%)** |  |  |
| T1a-c | 11521 (58.5) | 1753 (32.0) |
| T2a/b | 5749 (29.2) | 2748 (50.3) |
| T2c/T3a | 2132 (10.8) | 690 (12.6) |
| T3b/T4 | 282 (1.4) | 277 (5.1) |
| Unknown | 0 (0.0) | - |
| **Clinical nodal category (%)** |  |  |
| N0 | 19646 (99.8) | 5400 (98.8) |
| N1 | 38 (0.2) | 68 (1.2) |
| **Gleason grade (%)** |  |  |
| ⋦ 3+3 | 10153 (51.6) | 2861 (52.3) |
| 3+4 | 4897 (24.9) | 1957 (35.8) |
| 4+3 | 2405 (12.2) |  |
| 4+4 | 1411 (7.2) | 395 (7.2) |
| 4+5 | 593 (3.0) | 238 (4.4) |
| 5+4 | 160 (0.8) |  |
| 5+5 | 65 (0.3) | 17 ( 0.3) |
| **Biopsy cores** |  |  |
| Median sampled (IQR) | 10 (6, 12) | - |
| Median number positive (IQR) | 3 (1, 5) | - |
| Median percent positive (IQR) | 0.3 (0.2, 0.5) | - |
| **Prostate specific antigen (ng/mL)** |  |  |
| Median (IQR) | 6.3 (4.7, 9.5) | 6.0 (4.6, 8.7) |
| **Education (%)** |  |  |
| Less than 9th grade | - | 55 (1.0) |
| 9th-11th grade | - | 322 (5.9) |
| HS graduate | - | 1001 (18.3) |
| Some college | - | 1725 (31.5) |
| College graduate | - | 2365 (43.3) |
| **Marital status (%)** |  |  |
| Married | - | 4767 (87.2) |
| Separated | - | 567 (10.4) |
| Single | - | 134 (2.5) |
| **Smoking status (%)** |  |  |
| Never | - | 2227 (40.7) |
| Current | - | 467 (8.5) |
| Former | - | 2774 (50.7) |
| **Diabetes (%)** | - | 317 (5.8) |
| **Hypertension (%)** | - | 1760 (32.2) |
| **Previous stroke (%)** | - | 108 (2.0) |
| **Body mass index (%)** |  |  |
| < 18.5 | - | 12 (0.2) |
| 18.5-25 | - | 1511 (27.6) |
| 25-40 | - | 3907 (71.5) |
| 40+ | - | 38 (0.7) |
| **Primary treatment (%)** |  |  |
| RP | 12421 (63.1) | 2724 (49.8) |
| EBRT | 5942 (30.2) | 2744 (50.2)  -  - |
| BT | 1101 (5.6) |  |
| EBRT+BT | 220 (1.1) |  |
| **RT patients (%)** |  |  |
| RT alone | 3656 (50.3) | 1374 (50.0) |
| RT + short term ADT | 2178 (30.0) | - |
| RT + long term ADT | 696 (9.6) | - |
| RT + unknown ADT duration | 733 (10.1) | 1374 (50.0) |
| **Follow-up (months)** |  |  |
| Median (range) | 71.8 [0.0, 180.0] | 157 [0.0, 267.0] |
| **Outcome (%)** |  |  |
| Death from prostate cancer | 450 (2.3) | 244 (4.5) |
| Death from other causes | 2522 (12.8) | 1473 (26.9) |
| Alive | 16712 (84.9) | 3751 (68.6) |

**Table S3: Comorbidity profiles for low, median, and high OCM risk patients from the PLCO prostate cancer cohort.**

|  | **Age** | **Marital Status** | **Education** | **Smoking Status** | **BMI Category** | **Hypertension** | **Diabetes** | **Stroke** |
| --- | --- | --- | --- | --- | --- | --- | --- | --- |
| **Low Risk** | 71 | Married | Some college | Never | Overweight | No | No | No |
| **Median Risk** | 74 | Married | Some college | Former | Overweight | Yes | No | No |
| **High Risk** | 77 | Separated | Some college | Former | Overweight | Yes | Yes | No |

**Table S4: TRIPOD Guidelines Checklist**

| **Section (**¶s) | **Item** | **Checklist** | **Paragraph (**¶) |
| --- | --- | --- | --- |
| *Title and abstract* | | | |
| **Title** | **1** | *Identify the study as developing and/or validating a multivariable prediction model, the target population, and the outcome to be predicted.* | **Title** |
| **Abstract** | **2** | *Provide a summary of objectives, study design, setting, participants, sample size, predictors, outcome, statistical analysis, results, and conclusions.* | **Abstract** |
| **Introduction (**¶1-3) | | | |
| *Background*  *and objectives* | **3a** | *Explain the medical context (including whether diagnostic or prognostic) and rationale for developing or validating the multivariable prediction model, including references to existing models.* | ¶ 1-3 |
|  | **3a** | *Specify the objectives, including whether the study describes the development or validation of the model or both.* | ¶3 |
| **Methods (**¶4-9) | | | |
| *Source of data* | **4a** | *Describe the study design or source of data (e.g., randomized trial, cohort, or registry data), separately for the development and validation data sets, if applicable.* | ¶4; Supp |
|  | **4b** | *Specify the key study dates, including start of accrual; end of accrual; and, if applicable, end of follow-up.* | **Supp** |
| **Participants** | **5a** | *Specify key elements of the study setting (e.g., primary care, secondary care, general population) including number and location of centers.* | ¶4 |
|  | **5b** | *Describe eligibility criteria for participants.* | **Supp** |
|  | **5c** | *Give details of treatments received, if relevant.* | ¶4 |
| **Outcome** | **6a** | *Clearly define the outcome that is predicted by the prediction model, including how and when assessed.* | ¶5 |
|  | **6b** | *Report any actions to blind assessment of the outcome to be predicted.* | **NA** |
| **Predictors** | **7a** | *Clearly define all predictors used in developing or validating the multivariable prediction model, including how and when they were measured.* | ¶5-9,  **Supp** |
|  | **7b** | *Report any actions to blind assessment of predictors for the outcome and other Predictors.* | **NA** |
| **Sample Size** | **8** | *Explain how the study size was arrived at.* | **NA** |
| **Missing Data** | **9** | *Describe how missing data were handled (e.g., complete-case analysis, single imputation, multiple imputation) with details of any imputation method.* | **Supp** |
| **Statistical Analysis** | **10a** | *Describe how predictors were handled in the analyses.* | ¶5-9 |
|  | **10b** | *Specify type of model, all model-building procedures (including any predictor selection), and method for internal validation* | ¶5-9; Supp |
|  | **10d** | *Specify all measures used to assess model performance and, if relevant, to compare multiple models.* | ¶8-9; **Supp** |
| **Risk group** | **11** | *Provide details on how risk groups were created, if done.* | **NA** |
| **Results (**¶10-15) | | | |
| **Participants** | **13a** | *Describe the flow of participants through the study, including the number of participants with and without the outcome and, if applicable, a summary of the follow-up time. A diagram may be helpful.* | **Supp (Table S2)** |
|  | **13b** | *Describe the characteristics of the participants (basic demographics, clinical features, available predictors), including the number of participants with missing data for predictors and outcome.* | **Supp (Table S2)** |
| **Model Development** | **14a** | *Specify the number of participants and outcome events in each analysis.* | **Supp (Table S2)** |
|  | **14b** | *If done, report the unadjusted association between each candidate predictor and outcome.* | **NA** |
| **Model Specification** | **15a** | *Present the full prediction model to allow predictions for individuals (i.e., all regression coefficients, and model intercept or baseline survival at a given time point).* | **Supp** |
|  | **15b** | *Explain how to the use the prediction model.* | ¶5-9 |
| **Model Performance** | **16** | *Report performance measures (with CIs) for the prediction model* | ¶10-15 **Supp** |
| **Discussion(**¶16-19) | | | |
| **Limitations** | **18** | *Discuss any limitations of the study (such as nonrepresentative sample, few events per predictor, missing data).* | ¶18 |
| **Interpretation** | **19b** | *Give an overall interpretation of the results, considering objectives, limitations, and results from similar studies, and other relevant evidence.* | ¶16-19 |
| **Implications** | **20** | *Discuss the potential clinical use of the model and implications for future research.* | ¶16-19 |
| **Other Information** | | | |
| **Supplementary Information** | **21** | *Provide information about the availability of supplementary resources, such as study protocol, Web calculator, and data sets.* | **Supp** |
| **Funding** | **22** | *Give the source of funding and the role of the funders for the present study.* | **Disclosure** |

**Table S5. The Predictive Approaches to Treatment effect Heterogeneity (PATH) Consensus Criteria.**

| **Consensus Criteria** | **Section** |
| --- | --- |
| Overall treatment effect is well established | **Intro** |
| When benefits and harms/burden of treatment are finely balanced | **Intro** |
| Treatment associated with nontrivial amount of harm/burden | **Intro/Methods** |
| Several Large, well-conducted RCTs are available and appropriate to pool in individual patient meta-analyses | **Discussion (future work)** |
| Identifiable heterogeneity of risk in the trial population anticipated | **Intro/**  **Results** |
| Strong preliminary evidence prediction model is clinically useful for treatment selection | **Results** |
| Clinical variables in proposed model are routinely available | **Intro** |
